# Impact of combined medication payment management policies on population health performance

**DOI:** 10.1101/2025.08.01.25332645

**Authors:** Dingqiang Duan, Yun Yang

## Abstract

This study investigated the mechanism and impact of a policy combination involving centralized drug procurement and national drug price negotiations on health insurance payment management and overall health performance of the population. Relying on a two-dimensional analytical framework of health outcomes and medical expenditures, the entropy value method was applied to construct indicators of residents’ health portfolios. The year 2019, marked by the large-scale implementation of centralized procurement and national medicine catalog negotiations, was identified as the policy breakpoint for constructing a breakpoint regression model. Based on CFPS data, the model was implemented to evaluate changes in residents’ health performance and medical expenditure efficiency. Furthermore, the mechanisms underlying policy effects were examined from the perspectives of drug expenditure, pharmaceutical innovation (e.g., R&D inputs and patent output), and drug trade including imports and exports. The results indicated that this policy combination significantly improved population health outcomes and enhanced the efficiency of healthcare spending. The mechanism analysis further confirmed its short-term effects on stimulating innovation, increasing drug accessibility, and promoting expenditure efficiency. In addition, empirical evidence supported the hypothesized synergy between import substitution and export upgrading. Therefore, it is recommended to establish a value-oriented drug classification and payment management mechanism while adapting regional policies to provide a scientific basis for optimizing pharmaceutical policy design and balancing health accessibility with the advancement of innovation in the pharmaceutical industry.Keywords: Volume Procurement; national drug negotiation; health performance; breakpoint regression

## 1. Introduction

People’s health serves as a fundamental benchmark for evaluating the effectiveness of Chinese-style modernization. The 2025 Chinese Government Work Report emphasizes the implementation of a health-first development strategy to enhance the population’s well-being. Since 2018, the health insurance drug payment policy portfolio centered on volume-based procurement and the negotiation mechanism of the health insurance catalog has undergone systematic reform through the strategy of “vacating the cage and exchanging the bird”. This approach aims to rationalize drug prices, optimize the allocation of healthcare resources, and steer the pharmaceutical industry towards innovation and development through a dual mechanism of incentives and constraints [1]. According to statistics, the nationally organized volume-based procurement of drugs has cumulatively saved approximately 440 billion yuan in health insurance funds, of which over 360 billion yuan has been redirected to cover negotiated drugs. This reallocation, characterized by simultaneous cost reduction and quality improvement, has significantly enhanced the efficiency of health insurance fund utilization. The remaining savings have primarily supported public hospital salary reform. This comprehensive policy framework not only utilized price discovery to eliminate inflated generic drug prices but also leveraged strategic fund allocation to facilitate the inclusion of innovative drugs into the catalog. Additionally, salary system reforms have influenced the financial link between prescription behavior and rebate-driven incentives. Through this three-pronged strategy, the reductions in pharmaceutical expenditures, the elevation of medical service value, and the upgrading of industrial innovation capacity have been jointly achieved, realizing Pareto improvement and advancing the development of high-quality productivity in the pharmaceutical sector.

With the ongoing implementation of the Healthy China strategy, public expectations for accessibility, affordability, and quality of medical services continue to rise. However, despite the slow growth in national medical expenditure in recent years, the financial burden on individuals has increased and public concerns over the quality of rapidly substituted generic medicines have intensified. Therefore, it is of both theoretical and practical significance to conduct a scientific evaluation of whether the current medical insurance payment management policy combination enhances the efficiency of medical expenditure and improves health outcomes while also addressing public concerns and expectations.

In evaluating the performance of medication payment management policies, the existing literature has predominantly adopted a single-policy perspective, focusing on unidirectional impacts on the government [2], hospitals [3], and pharmaceutical companies [4]. However, such studies often lack a comprehensive assessment of policy combinations and fail to decompose and examine policy outcomes in terms of both performance and effectiveness. Consequently, these limitations hinder the capacity to fully address the objectives of the “Healthy China” strategy, aimed at enhancing public well-being and health. In response, this study developed a theoretical analytical framework of “policy tool-market response-health performance” and employed the entropy weight method to construct composite indicators reflecting both the “performance” and “effectiveness” of residents’ health outcomes. Furthermore, a regression analysis was applied to evaluate the outcomes of the integrated policy approach under the “Healthy China” strategy. Utilizing a breakpoint regression model and CFPS microdata, this study assessed the influence of these policies on residents’ health outcomes and economic burden, while further analyzing the underlying mechanisms through factors such as residents’ pharmaceutical expenditures, pharmaceutical innovation (including R&D investment, patent output, and import-export activity of medicines), among others.

The key innovations of this study are as follows. First, a two-dimensional evaluation system of “health benefits and healthcare expenditure efficiency” was analyzed to examine how policy combinations could improve health outcomes by reshaping the competitive structure of the pharmaceutical market and encouraging innovation. Second, it moved beyond the limitations of traditional closed-system evaluations, revealing the mechanisms of domestic innovation incentives (supply side) and global value chain embeddedness (distribution side) in shaping health outcomes. These findings provide new theoretical foundations for designing pharmaceutical payment policies in an open and competitive market environment.

## 2. Theoretical Analysis and Research Hypotheses

### 2.1 Impact of Medicare Medication Payment Management Policies on Residents’ Health Performance

Based on the concept of value-based healthcare, this study established a dual-dimensional framework for evaluating policy performance. The first dimension concerned residents’ physical health benefits, encompassing physiological functions, disease conditions, and quality of life, to represent the core objective of health policy. The second dimension focused on the efficiency of medical expenditure, reflecting the comprehensive nature of healthcare services, including accessibility, drug quality, and rational drug use.

#### 2.1.1 Drug price reduction affects patients’ medication behavior

Drug prices are a critical factor influencing patient medication decisions. Payment management policies that reduce prices contribute to improved medication adherence. Specifically, the inclusion of drugs in centralized volume-based procurement and national negotiation lists [5] significantly lowers prices, thereby easing the economic burden on patients, conserving medical insurance funds [6], and improving health outcomes [7]. Simultaneously, reduced drug prices can enhance patients’ adherence to treatment, minimizing the likelihood of medication reduction or discontinuation due to financial constraints [8] and thereby preventing deterioration in health status. Nevertheless, lower costs may also trigger an excessive release of latent “health needs”, potentially resulting in overutilization of low-value medical services [9] and diminished engagement in preventive health behaviors [10].

#### 2.1.2 Differences in accessibility of different categories of drugs due to centralized purchasing and health insurance catalogs

The combination of centralized procurement and health insurance catalog negotiations improves drug accessibility by leveraging scale effects. The policy facilitates “exchange volume for price” arrangements [11], resolving issues such as the decoupling of price and volume, weak bargaining power in decentralized procurement, and lack of policy cohesion. Although such mechanisms enhance public welfare through price transfers [12], excessive price suppression may diminish pharmaceutical companies’ incentives for R&D investment [13], potentially compromising the long-term quality and availability of innovative drugs [14]. Moreover, inadequate provision of innovative therapies in primary care coupled with hospitals’ preference for low-priced options may result in insufficient access to high-quality drugs, thereby limiting the actual improvement in drug affordability and negatively affecting patient outcomes.

#### 2.1.3 Access gap under policy effectiveness

The policy has yielded preliminary outcomes in curbing medical expenses by reducing drug distribution costs [15]. The composition of hospitalization expenditures has been optimized, with a marked decline in the proportion allocated to medications, thereby providing institutional support for alleviating patients’ financial burden related to pharmaceuticals [16]. However, a discrepancy remains between the observed reduction in out-of-pocket expenses and the actual perception of access among residents. On the one hand, the adjustments to payment standards for non-negotiated drugs have limited access. On the other hand, price reductions have released previously suppressed demand [17]. This mismatch between macro-level cost control and micro-level access perception has led to divergence between the policy’s intended outcomes and residents’ experiences.

Hypothesis 1: The combination of medication payment management policies improves the population health performance.

### 2.2 Pharmaceutical innovation and health performance

China’s health insurance drug payment management policy has a dual impact on the pharmaceutical innovation ecosystem by reshaping the market incentive structure. From the perspective of the domestic market, in response to the biopharmaceutical industry’s long-standing emphasis on marketing over R&D [18], volume-based procurement policy has compelled enterprises to transition towards innovation-driven models by constraining the marketing scope of generic drugs and reducing the market share of originator drugs with expired patents [19]. However, the policy’s incentive effect on innovation remains limited. Acceleration of access to innovative drugs and the provision of payment protection through health insurance negotiations have been designed to establish clearer innovation return expectations, thereby promoting a gradual transition of the industry toward innovation-driven development.

In the international value chain dimension, policies have facilitated domestic substitution by lowering the prices of imported originator drugs [20]. Nonetheless, markets in technology-intensive fields and rare disease treatment remain heavily reliant on imports. While such imports help address accessibility issues for patented drugs, multinational pharmaceutical corporations continue to reap excessive profits through patent monopolies [21], intensifying the medication burden in developing countries. In response, the Chinese government has implemented price controls and expanded national drug negotiations to reduce the economic burden on patients and improve therapeutic effectiveness [22]. This has led to the formation of a synergistic policy path of “import substitution-export upgrading”. This policy-induced dual mechanism linking local innovation incentives with global value chain restructuring reflects the internal logic of reforms in the drug payment management system. It enhances health performance by optimizing the allocation of innovation-related resources.

Hypothesis 2: The health insurance medication payment policy mix enhances population health performance by reducing residents’ drug expenditures, incentivizing pharmaceutical firms’ R&D innovation, and leveraging synergies from import substitution and export upgrading within global value chains.

## 3 Research Design

### 3.1 Resident Health Performance Indicators and Measurement

#### 3.1.1 Indicator Design

Building on prior analysis, this study developed a two-dimensional indicator system for evaluating residents’ health performance. The system integrates health perception, disease risk, and medical expenditure to comprehensively assess both health benefits and expenditure efficiency. The indicators were derived from CFPS survey questions. Weights were assigned to sub-indicators using the entropy weight method, and quantitative measures were calculated based on **de-identified public data from the China Family Panel Studies (CFPS) during 2016-2022 【 accessed in January 2025】.

##### Ethical Compliance

This research complies with all ethical standards for secondary data analysis: The CFPS project was approved by the Biomedical Ethics Committee of Peking University (IRB No. IRB00001052-14010). Written informed consent was obtained from all original participants:

- Respondents aged ≥16 years signed independently.
- Respondents aged ≤15 years provided consent through guardians.

- 【Data were accessed in January 2025 】 from fully anonymized repositories with no identifiable information.

(Consent form template: [CFPS Website](http://www.isss.pku.edu.cn/cfps/news/index.htm))

**Table 1.**
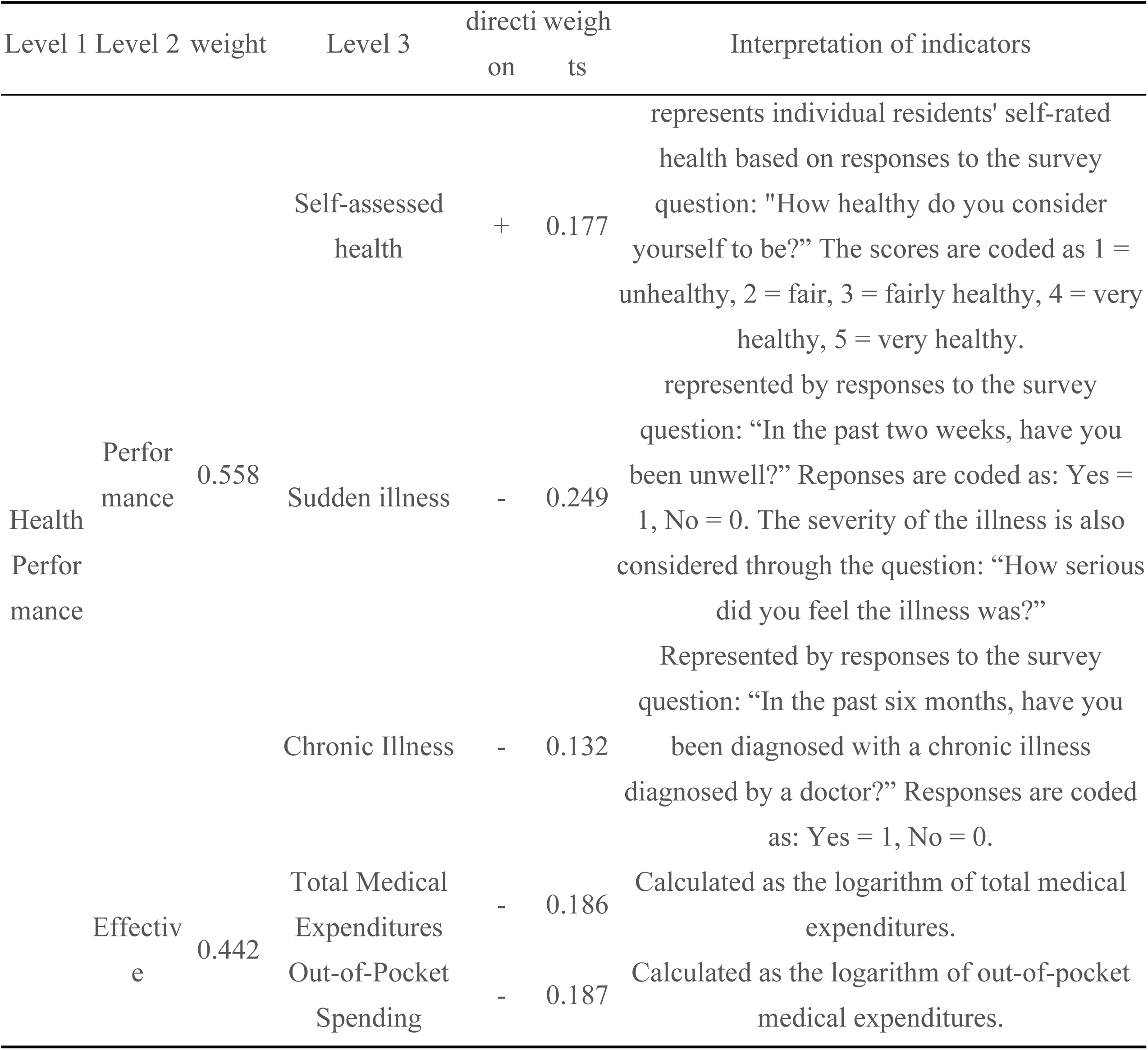
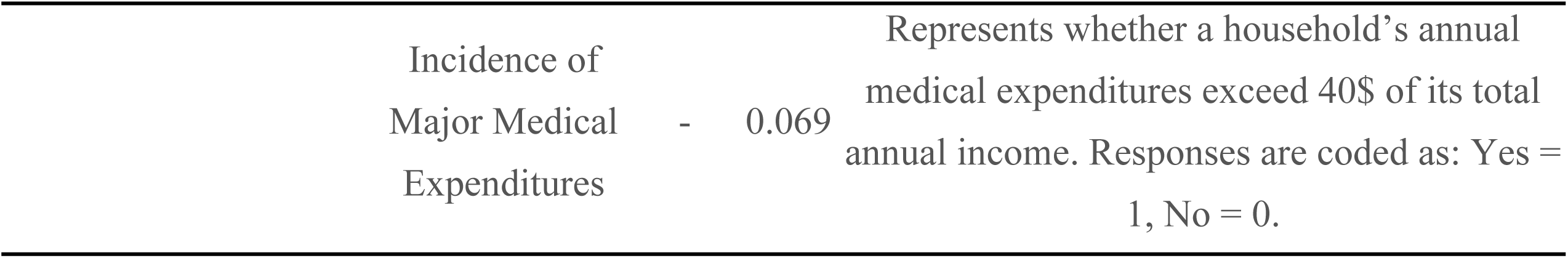
Design and weights of health performance evaluation indicator system.

#### 3.1.2. Analysis of measurement results

The data presented in Table 2 indicate that residents’ health performance in China has undergone a notable upward trend and structural transformation. From 2016 to 2022, the average annual growth rate of overall health performance reached 1.2%, with a particularly marked increase from 2018 to 2020. From a compositional perspective, the health status component exhibited a steady improvement, with an average annual growth rate of 0.6%. Meanwhile, the efficiency component, represented by medical expenditure indicators, demonstrated a more rapid increase, averaging 1.8% annually, which was approximately three times the rate of health status improvement. This trend underscores the strong and immediate impact of health insurance payment reform on medical expenditure efficiency.

**Table 2.**
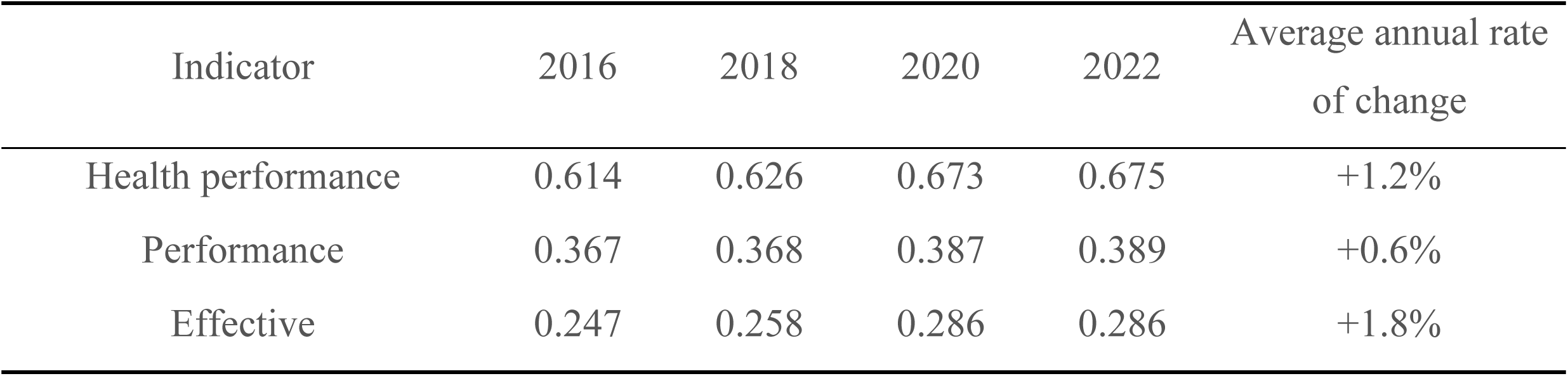
Mean scores of national population health performance.

| Indicator | 2016 | 2018 | 2020 | 2022 | Average annual rate<br>of change |
| --- | --- | --- | --- | --- | --- |
| Health performance | 0.614 | 0.626 | 0.673 | 0.675 | +1.2% |
| Performance | 0.367 | 0.368 | 0.387 | 0.389 | +0.6% |
| Effective | 0.247 | 0.258 | 0.286 | 0.286 | +1.8% |

As shown in Figure 1, the national and regional health performance trends from 2016 to 2022 were largely consistent. All regions experienced an initial decline from 2016 to 2018, followed by a marked post-policy increase after the 2019 implementation, and a stabilization phase between 2020 and 2022. Regionally, Eastern and Central China consistently outperformed the national average, while Western China remained below the national level. Notably, the Western region showed the most significant improvements during the policy period. Despite regional differences in baseline performance, all areas demonstrated measurable enhancements in health performance driven by policy intervention. This improvement was accompanied by a gradual narrowing of regional disparities over time.

**Figure 1.**
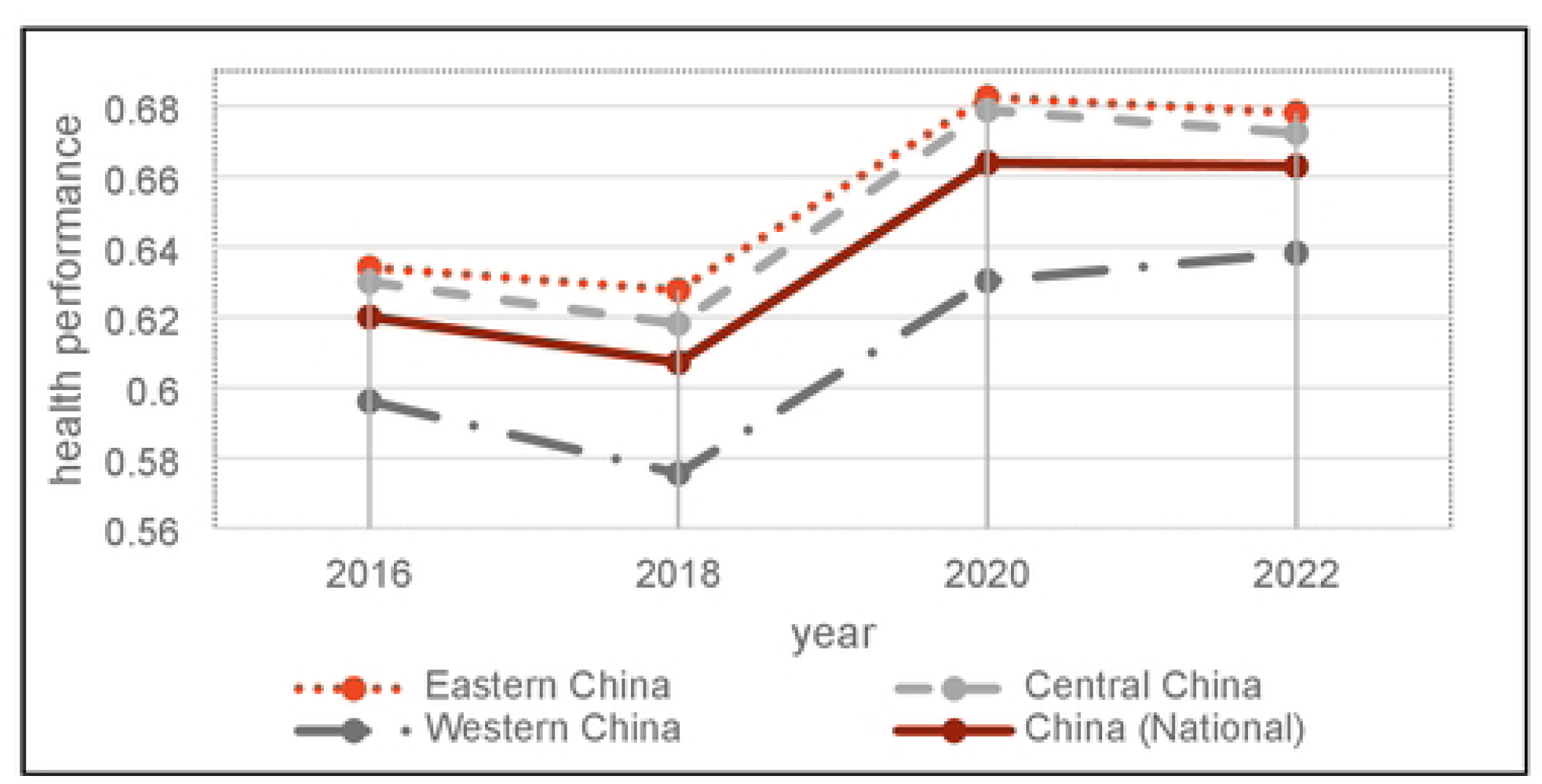
Trends in population health performance at the national level and across the eastern, central, and western regions, 2016–2022.

Figure 2 tracks the evolution of population health sub-indicators, namely “performance” and “effectiveness”. While performance indicators follow similar trajectories across Eastern, Central, and Western China, persistent regional disparities remain throughout the study period, with no signs of convergence. In contrast, effectiveness indicators reflecting differences in regional health expenditure levels display significant initial variations. However, after the implementation of the health insurance medication payment policies, the gap in effectiveness across regions began to narrow, suggesting a convergence trend over time.

**Figure 2.**
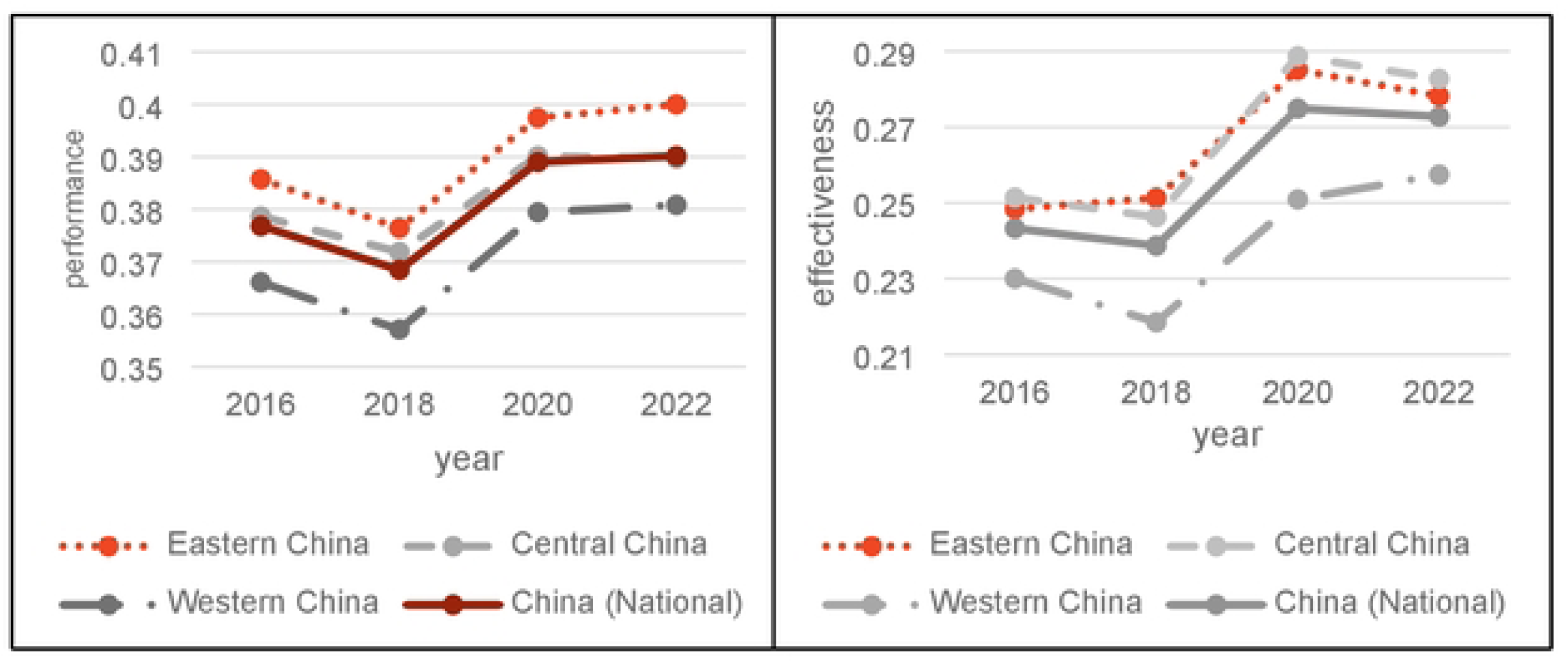
Trends in population “performance” and “effectiveness” at the national level and across the eastern, central, and western regions, 2016–2022.

### 3.2 Empirical Model Setting

#### 3.2.1 Time Breakpoint Regression Model

To estimate the causal impact of health insurance medication payment policies on population health outcomes, this study employed a Regression Discontinuity in Time (RDiT) model, originally proposed by Hausman and Rapson (2018) [23]. This model was designed to accommodate time-series data by using the timing of policy intervention as the discontinuity point. By segmenting the time axis and observing whether key outcome variables changed abruptly around the policy breakpoint, the model enabled causal inference between the policy and its effects.

Given that the policy was implemented uniformly and significantly across provinces and cities, the chosen breakpoint for this study was the year in which the policy was launched. Specifically, January 2019 when volume-based procurement reform and national drug catalog negotiations were officially rolled out was used as the breakpoint. Subsequent empirical analysis defined the treatment variables based on this temporal marker.

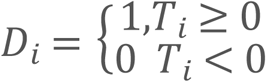

where T_i is the driving variable, indicating the monthly interval between the time and the “tipping point” of the policy implementation; T_i ≥ 0 indicates that the time is after January 2019; T_i < 0 indicates that the time is before January 2019; and D_i is the treatment variable, indicating whether provinces and municipalities in China are affected by the policy reform.

The average treatment effect (ATE) of the policy is defined as the difference in the expected value of the sample’s dependent variable immediately before and after the breakpoint. The model specification is as follows:

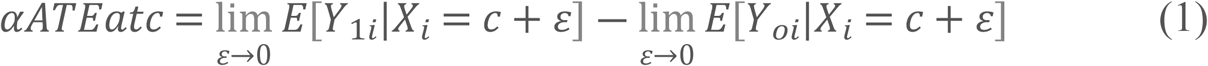

where αATEatc is the average treatment effect; Y_oi is the control group individuals before the break date; and Y_1i is the individuals treated by the policy after the break date.

The panel model was set up as follows:

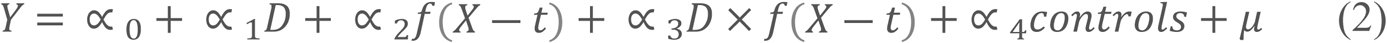

where Y is the health performance indicator, D is a treatment variable for the Medicare medication payment management policy mix, which takes the value of 0 before the policy is implemented and the value of 1 after the policy is implemented; x is a driver variable indicating the month in which the surveyed resident is located; f(x-t) is a higher-order polynomial of (x); controls is a control variable; and μ is a random perturbation term.

### 3.3 Data sources and variable definitions

#### 3.3.1 Data Sources

The data used in this study were sourced primarily from CFPS conducted by the China Center for Social Science Surveys at Peking University. Micro-level panel data from four CFPS waves (2016–2022) were used. Due to extensive missing data on children’s health indicators, the analysis was restricted to insured individuals aged 16 and above. Data on individual medication expenditure were obtained from the China Health and Wellness Statistical Yearbook. The firm-level variables were extracted from the Wind database, whereas the drug import and export data were derived from the General Administration of Customs of China.

#### 3.3.2 Variable Definition

**Table 3.**
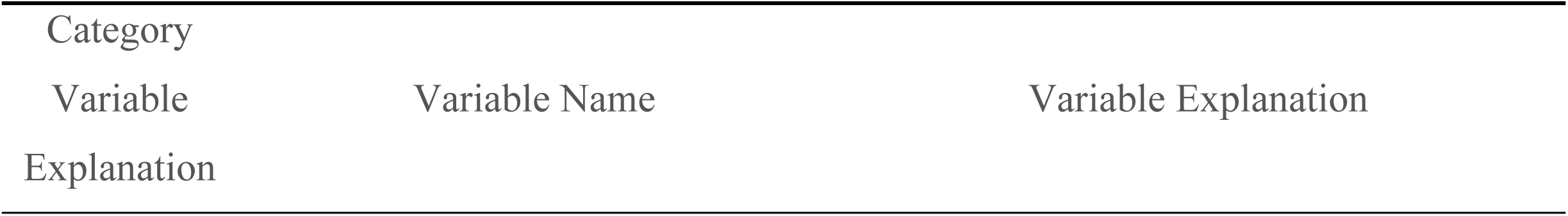
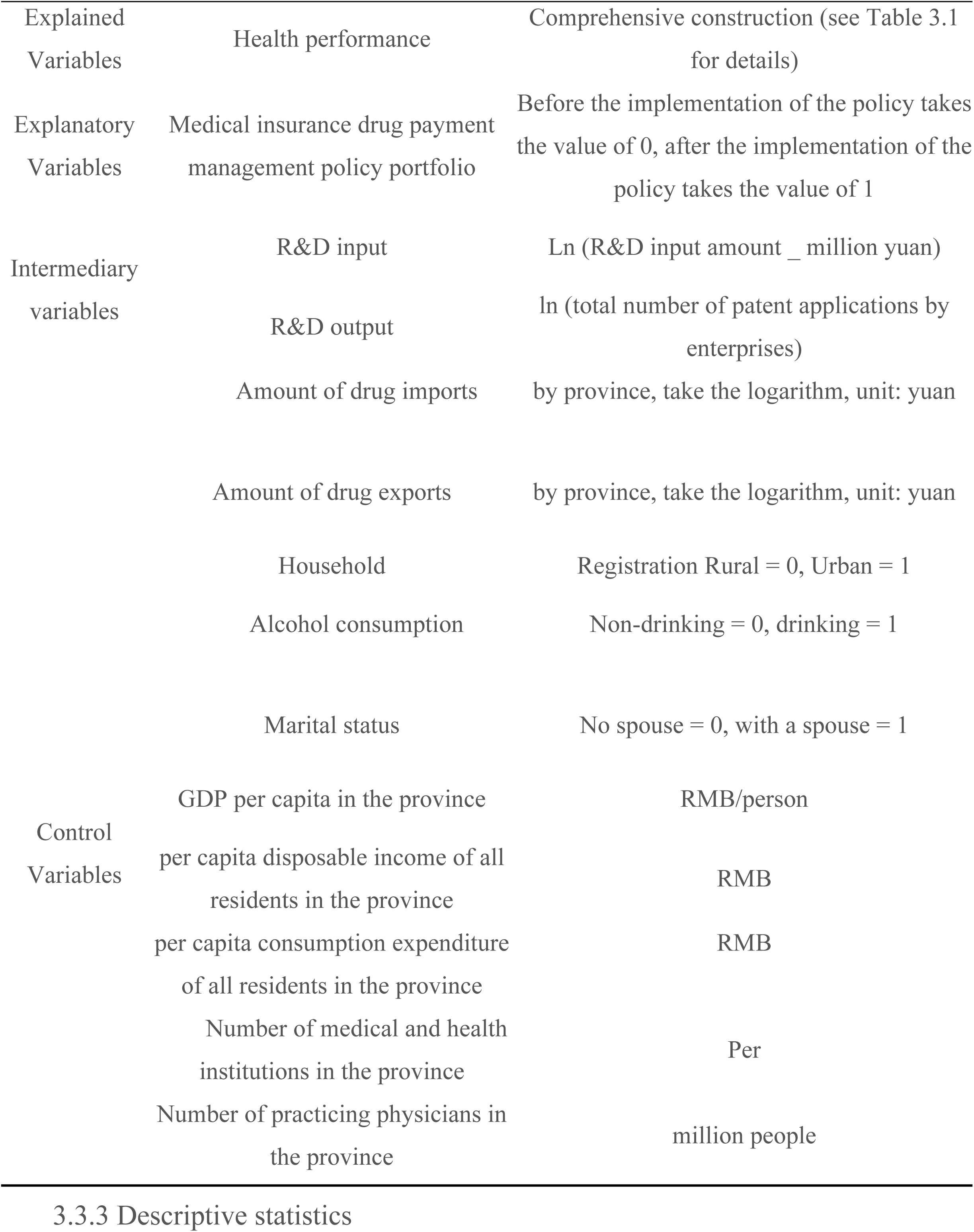
Definition of variables.

#### 3.3.3 Descriptive statistics

Table 4 presents the descriptive statistics for the variables in Table 3.3.

**Table 3.4.**
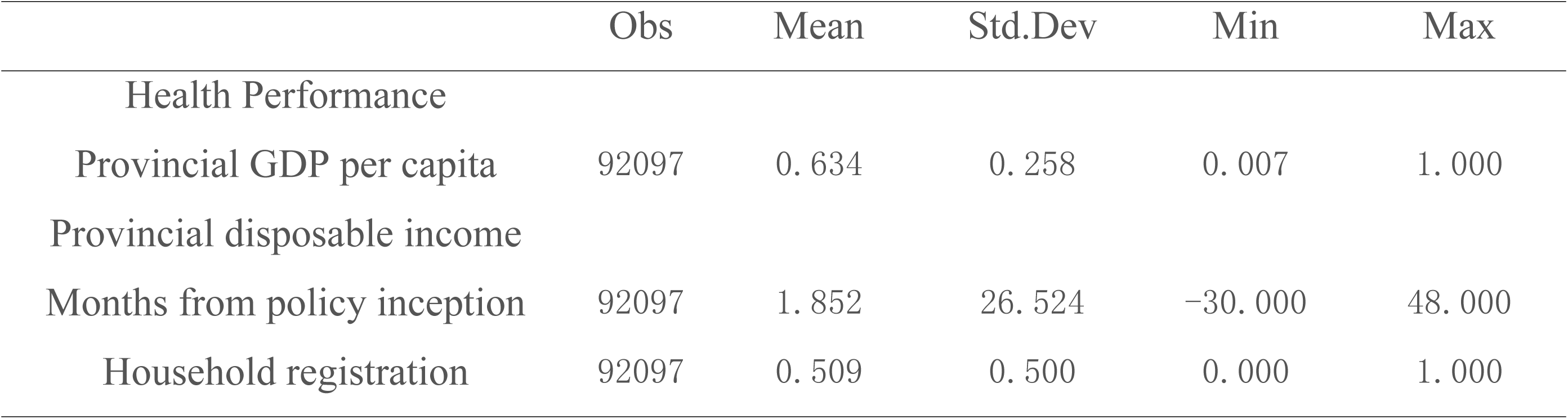
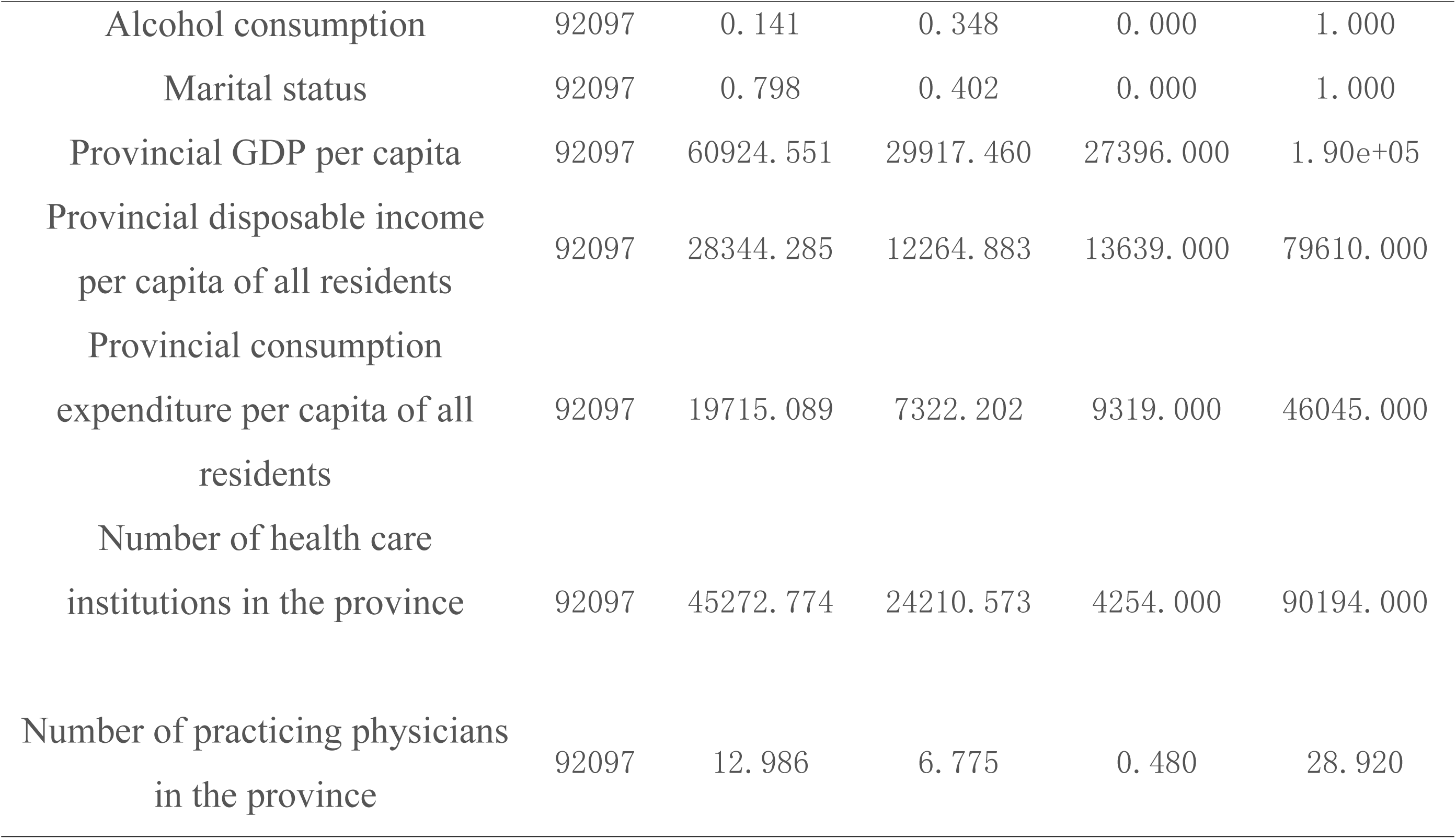
Descriptive statistics.

|  | Obs | Mean | Std.Dev | Min | Max |
| --- | --- | --- | --- | --- | --- |
| Health Performance |  |  |  |  |  |
| Provincial GDP per capita | 92097 | 0.634 | 0.258 | 0.007 | 1.000 |
| Provincial disposable income |  |  |  |  |  |
| Months from policy inception | 92097 | 1.852 | 26.524 | -30.000 | 48.000 |
| Household registration | 92097 | 0.509 | 0.500 | 0.000 | 1.000 |
| Alcohol consumption | 92097 | 0.141 | 0.348 | 0.000 | 1.000 |
| Marital status | 92097 | 0.798 | 0.402 | 0.000 | 1.000 |
| Provincial GDP per capita | 92097 | 60924.551 | 29917.460 | 27396.000 | 1.90e+05 |
| Provincial disposable income<br>per capita of all residents | 92097 | 28344.285 | 12264.883 | 13639.000 | 79610.000 |
| Provincial consumption<br>expenditure per capita of all<br>residents | 92097 | 19715.089 | 7322.202 | 9319.000 | 46045.000 |
| Number of health care<br>institutions in the province | 92097 | 45272.774 | 24210.573 | 4254.000 | 90194.000 |
| Number of practicing physicians<br>in the province | 92097 | 12.986 | 6.775 | 0.480 | 28.920 |

## 4. Empirical analysis

### 4.1 Breakpoint regression premise hypothesis testing

Breakpoint regression relies on the satisfaction of the three key conditions. First, the running variable should be exogenous. This was tested using McCrary’s density test. Following Gao and Pack [24], the running variable in this study was calendar time, which was inherently exogenous within the framework of time-based breakpoint regression. The assignment of time was determined objectively by year and was not subject to the residents’ subjective choices. Therefore, manipulation around the policy breakpoint was unlikely to satisfy the first condition. Second, discontinuity should exist in the outcome variable at the breakpoint. This was tested by fitting regression lines on either side of the breakpoint. As shown in Figure3, a significant upward shift in population health performance was observed at the breakpoint, confirming that the second condition was fulfilled.

**Figure 3.**
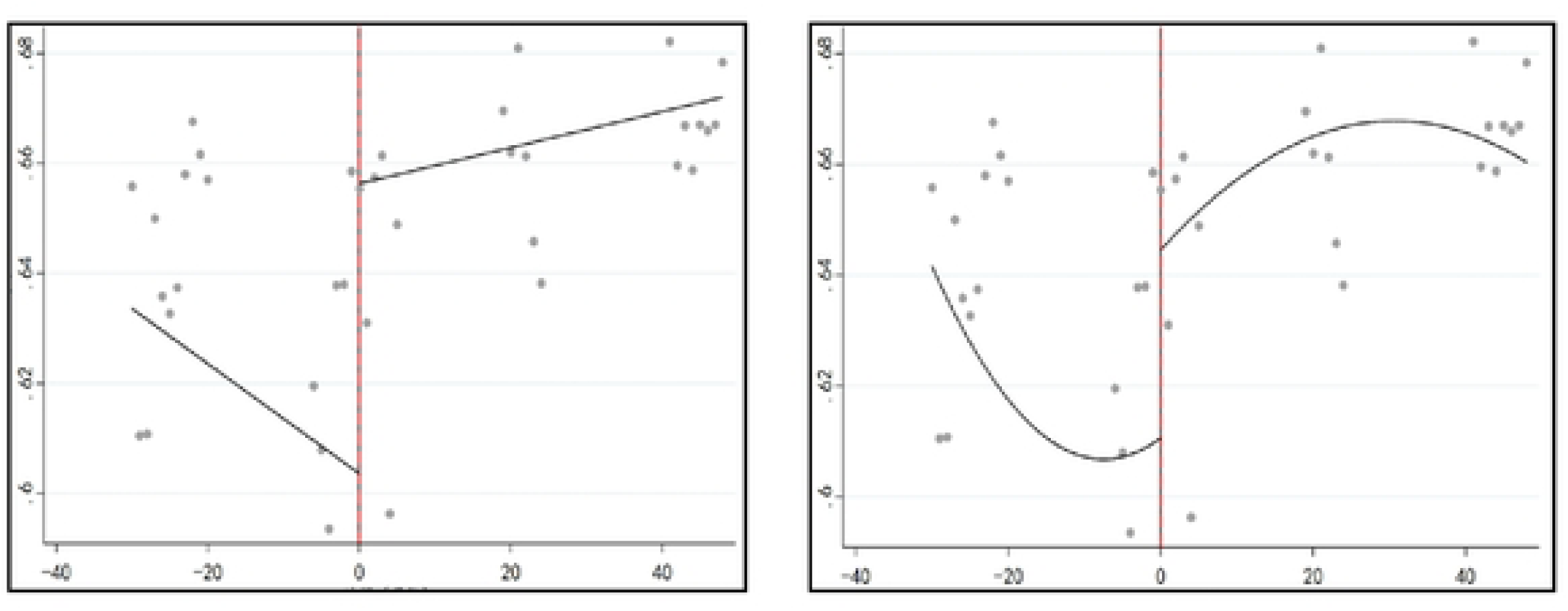
Changes in population health performance before and after breakpoints (left: primary fit; right: secondary fit).

Third, no discontinuity should occur in the covariates at the breakpoint. Table 5 presents the results of placebo tests using covariates as the outcome variables. These results showed no significant jumps, thereby satisfying the third condition.

**Table 5.**
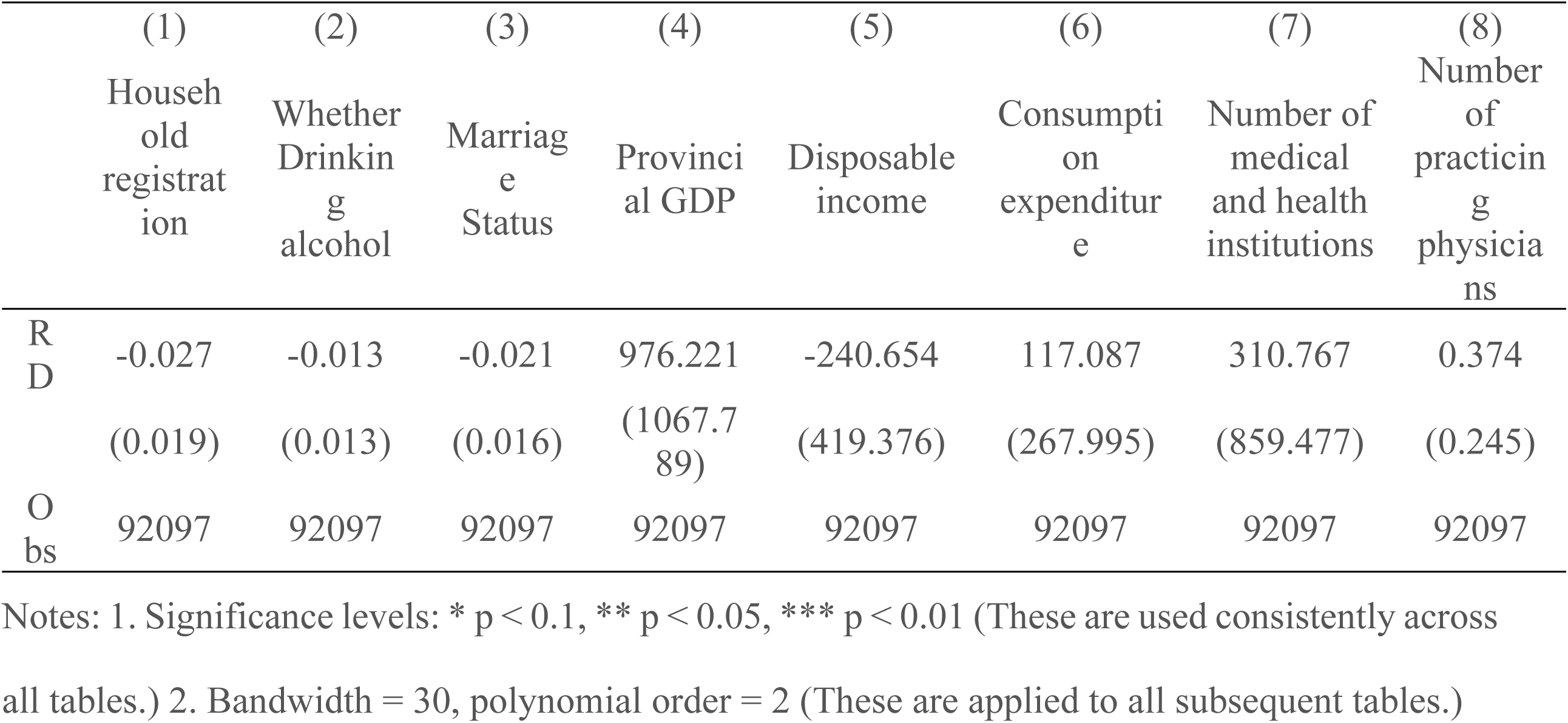
Covariate continuity tests for breakpoint regression.

### 4.2 Regression results

Based on the established indicator system, the regression analyses were conducted separately for “health performance”, “performance”, and “effectiveness”. The estimation results presented in Tables 6 to 7 indicated that policy intervention exerted a significant positive impact on population health performance. From a dimensional perspective, the policy demonstrated heterogeneous effects. After controlling for province fixed effects, linear time trends, and relevant covariates, the overall health performance of residents increased by 3.2% at the 1% significance level. Specifically, performance improved by 1.1%, while effectiveness increased by 2.4%, suggesting a more pronounced enhancement in expenditure-related efficiency than in health status.

**Table 6.** Breakpoint regression estimation of population health performance.

|  | (1) | (2) | (3) |
| --- | --- | --- | --- |
|  | Health performance |  |  |
| RD | 0.032***<br>(0.009) | 0.041***<br>(0.010) | 0.032***<br>(0.010) |
| Control variables | No | No | Yes |
| Province fixed effects | No | Yes | Yes |
| Linear time trend | No | Yes | Yes |
| Obs | 92,097 | 92,097 | 92,097 |

**Table 7.**
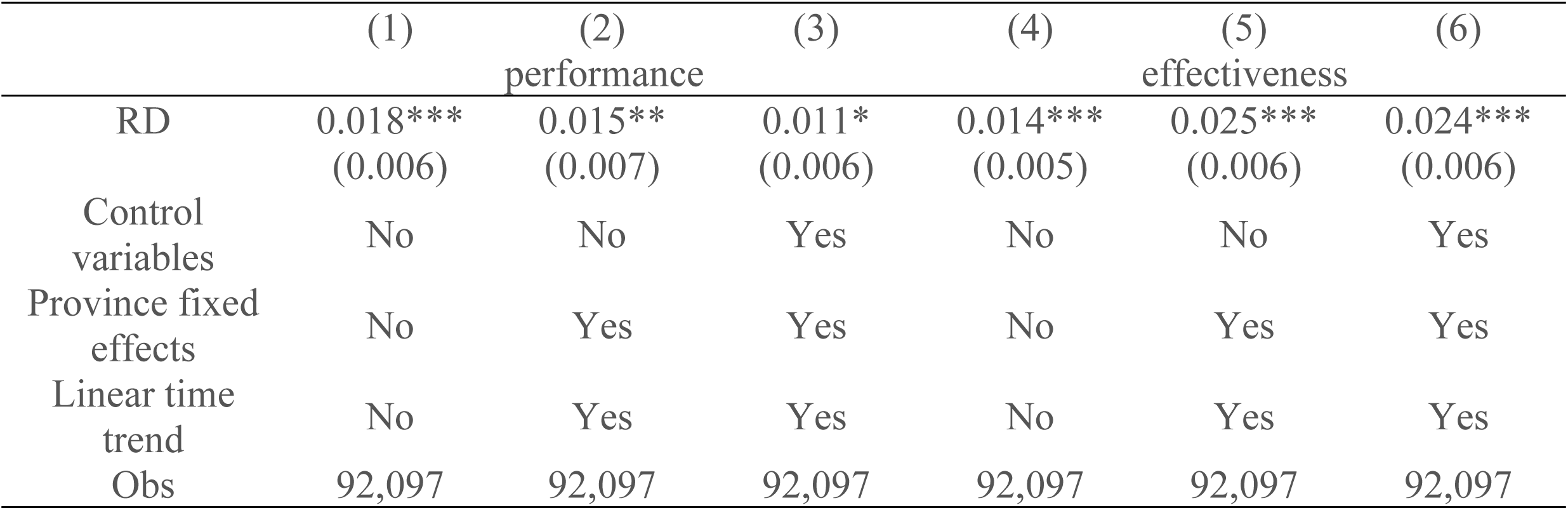
Breakpoint regression estimation of “performance” and “effectiveness” on population health.

To further evaluate the specific policy impacts, breakpoint regressions were performed on the individual “performance” and “effectiveness” indicators (Table 8). After policy implementation, the health metrics related to “performance” exhibited statistically significant improvements. Meanwhile, the “effectiveness” indicators demonstrated a substantial reduction in total medical expenditure. However, the reduction in out-of-pocket costs was not statistically significant. These findings suggested that the policy effectively promoted the intensive allocation of medical resources by regulating drug prices and improving reimbursement mechanisms, thereby confirming Hypothesis 1. The empirical results validated the effectiveness of the policy in enhancing health performance and optimizing medical resource allocation. However, these findings did not confirm a significant reduction in the economic burden on individual patients, indicating the need for continued attention from policymakers to address this aspect.

**Table 8.** Regression results for component indicators of population health performance.

|  | (1)<br>Self-<br>assessment<br>Level of<br>health | (2)<br>Sudden<br>illness | (3)<br>Chronic<br>illness | (4)<br>Total<br>medical<br>expenses | (5)<br>Out-of-pocket<br>spending | (6)<br>Incidence<br>of major<br>medical<br>expenses |
| --- | --- | --- | --- | --- | --- | --- |
| RD | 0.196***<br>(0.043) | -0.012<br>(0.017) | -0.044***<br>(0.012) | -616.958*<br>(327.895) | -146.586<br>(214.644) | -0.032***<br>(0.009) |
| Obs | 92,097 | 92,097 | 92,097 | 92,096 | 92,097 | 92,097 |

### 4.3 Analysis of regional heterogeneity

Table 9 presents the region-specific impacts of the policy on health performance indicators. In Eastern China, although the indicators are positive, they are statistically insignificant, suggesting a marginal effect saturation due to already mature healthcare systems. Central China shows positive but insignificant results in both composite and performance indicators, while effectiveness indicators display a significant improvement, indicating that the policy has effectively driven medical expenditure reduction in this region. In contrast, Western China exhibits significant positive changes across all dimensions. These improvements are attributed to the enhanced accessibility of primary care and increased availability of medications in this resource-constrained region. The findings suggest that the policy combination is most effective in improving health performance, where baseline medical resources are the weakest. Meanwhile, marginal returns diminish in more developed eastern provinces, and expenditure-oriented benefits are more pronounced in the central regions.

**Table 9.** Separate regression results for population health performance by region.

|  | (1) | (2) | (3) | (4) | (5) | (6) |
| --- | --- | --- | --- | --- | --- | --- |
|  | Eastern Region |  |  | Central Region |  |  |

|  | Health<br>Performanc<br>e | Performanc<br>e | effectivenes<br>s | Health<br>Performanc<br>e | Performanc<br>e | effectivenes<br>s |
| --- | --- | --- | --- | --- | --- | --- |
| RD | 0.011<br>(0.016) | 0.007<br>(0.011) | 0.004<br>(0.009) | 0.020<br>(0.016) | 0.002<br>(0.010) | 0.019**<br>(0.009) |
| Obs | 29,661 | 29,661 | 29,661 | 34,707 | 34,707 | 34,707 |

|  | (7) | (8) | (9) |
| --- | --- | --- | --- |
|  | Western Region |  |  |
|  | Health Performance | Performance | effectiveness |
| RD | 0.058***<br>(0.015) | 0.040***<br>(0.010) | 0.018**<br>(0.009) |
| Obs | 27,729 | 27,729 | 27,729 |

## 5. Robustness test

### 5.1 Bandwidth Selection Sensitivity Test

Bandwidth selection is a critical factor that influences the unbiasedness and validity of the breakpoint regression model. By estimating the model across different bandwidth settings, the effect of the intervention can be evaluated over various time horizons. As shown in Table 10 (Models 1 and 2), the core conclusions remain robust when alternative bandwidths are applied.

### 5.2 Kernel function selection sensitivity test

The selection of kernel function affects the precision of local average treatment effect (LATE) estimation by altering the assignment of weights to observations near the breakpoint. Table 10 (Models 3 and 4) demonstrates that the main findings remained consistent when different kernel functions such as Epanechnikov and Uniform were used, indicating the robustness of the model specification.

**Table 10.** Regression results for different bandwidths and kernel function breakpoints.

|  | (1)<br>h=36 | (2)<br>h=42 | (3)<br>Epanechnikov | (4)<br>Uniform |
| --- | --- | --- | --- | --- |
| RD | 0.040***<br>(0.008) | 0.050***<br>(0.009) | 0.043***<br>(0.009) | 0.060***<br>(0.009) |
| Obs | 92,097 | 92,097 | 92,097 | 92,097 |

### 5.3 Order Choice Sensitivity Tests

Model (1) in Table 11 shows that the breakpoint estimates remained significantly positive when a first-order polynomial was used. This finding confirmed that health performance improved even under different polynomial order assumptions, thereby supporting the robustness of the policy effectiveness hypothesis.

### 5.4 Placebo test

Placebo tests based on hypothetical policy shocks (January 2017 and January 2021) were conducted to ensure the credibility of the 2019 policy effect. The results of local linear regressions across these artificial breakpoints (Table 11, Models 2 and 3) revealed statistically insignificant estimates, confirming that the observed effects were not driven by spurious correlations. These findings validated the causal effect of the actual policy intervention in 2019.

**Table 11.**
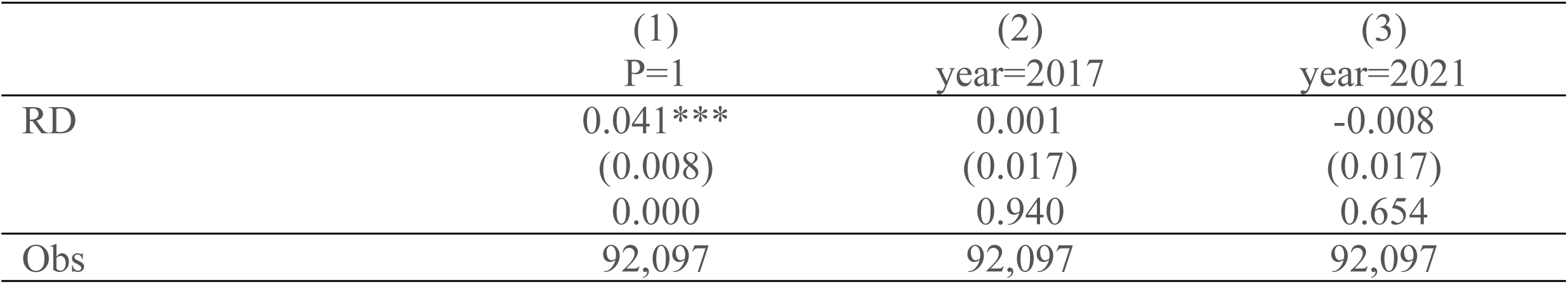
Results of different orders and placebo tests.

## 6. Discussion of drug prices and mechanisms of action that affect quality

Focusing on volume-based procurement and national drug negotiations, China’s medication payment policies utilize pricing mechanisms to reshape physician and patient behavior. This is reflected in the shift in prescription patterns toward innovative and generic drugs, resulting in reduced medication costs and accelerated pharmaceutical innovation. This study established a unified framework that integrates medication expenditures and drug innovation to evaluate the policy’s impact on population health performance. The analysis first investigated how drug price reductions influence health outcomes by generating cost savings for patients. Subsequently, this study explored policy-induced innovation effects by synthesizing R&D data and import-export indicators across domestic and international markets. This dual-pathway analysis revealed the mechanisms by which payment reforms improved both healthcare efficiency and health outcomes.

### 6.1 Expenditures on medicines for the population

Using provincial-level medication cost data (2016–2022) from the China Health Statistical Yearbook, combined with household survey–based health indicators, a breakpoint regression was conducted to estimate the impact of medication payment policies. Fixed-effects models were further applied to assess the influence of outpatient drug expenditure on health outcomes.

**Figure 4.**
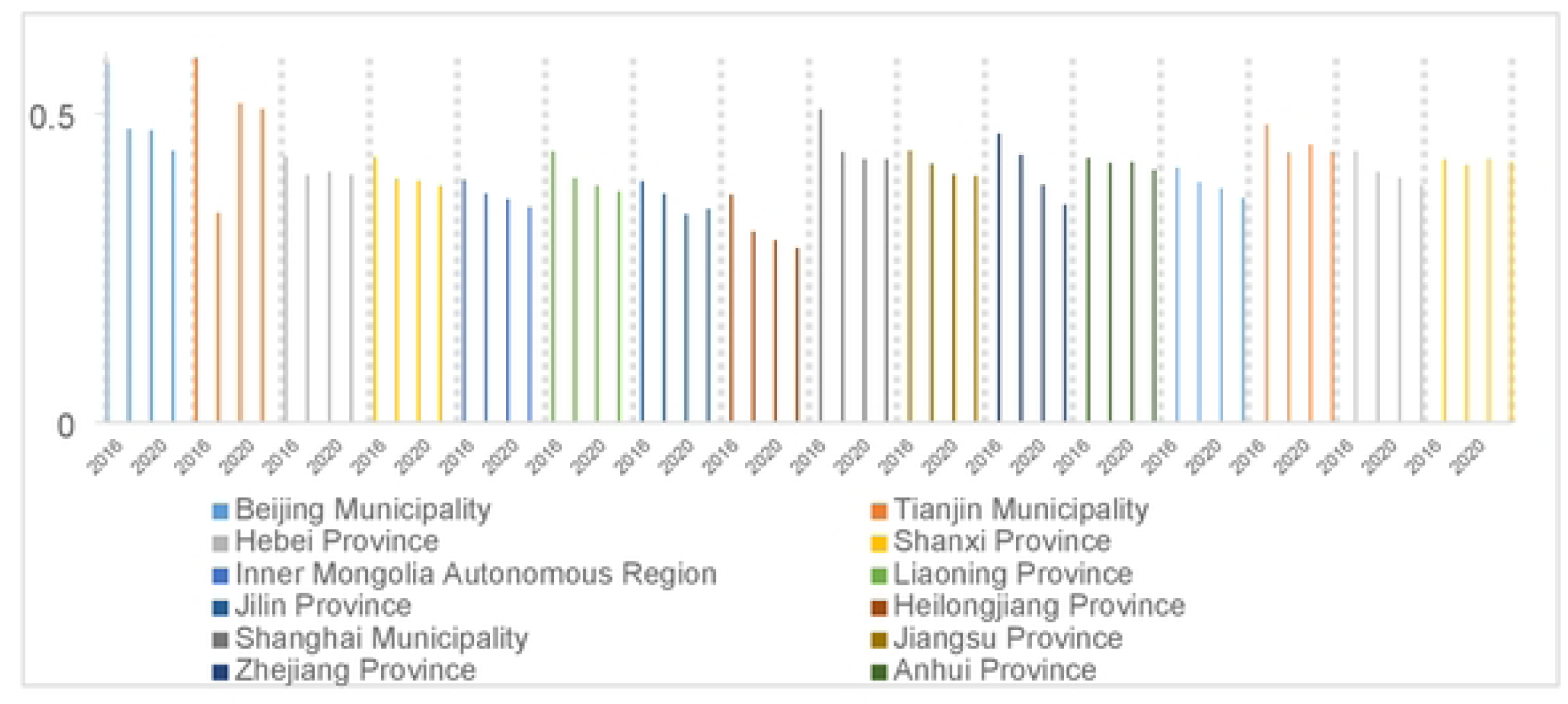
Average outpatient drug costs per visit as a percentage of total expenditures, 2016–2022.

**Figure 5.**
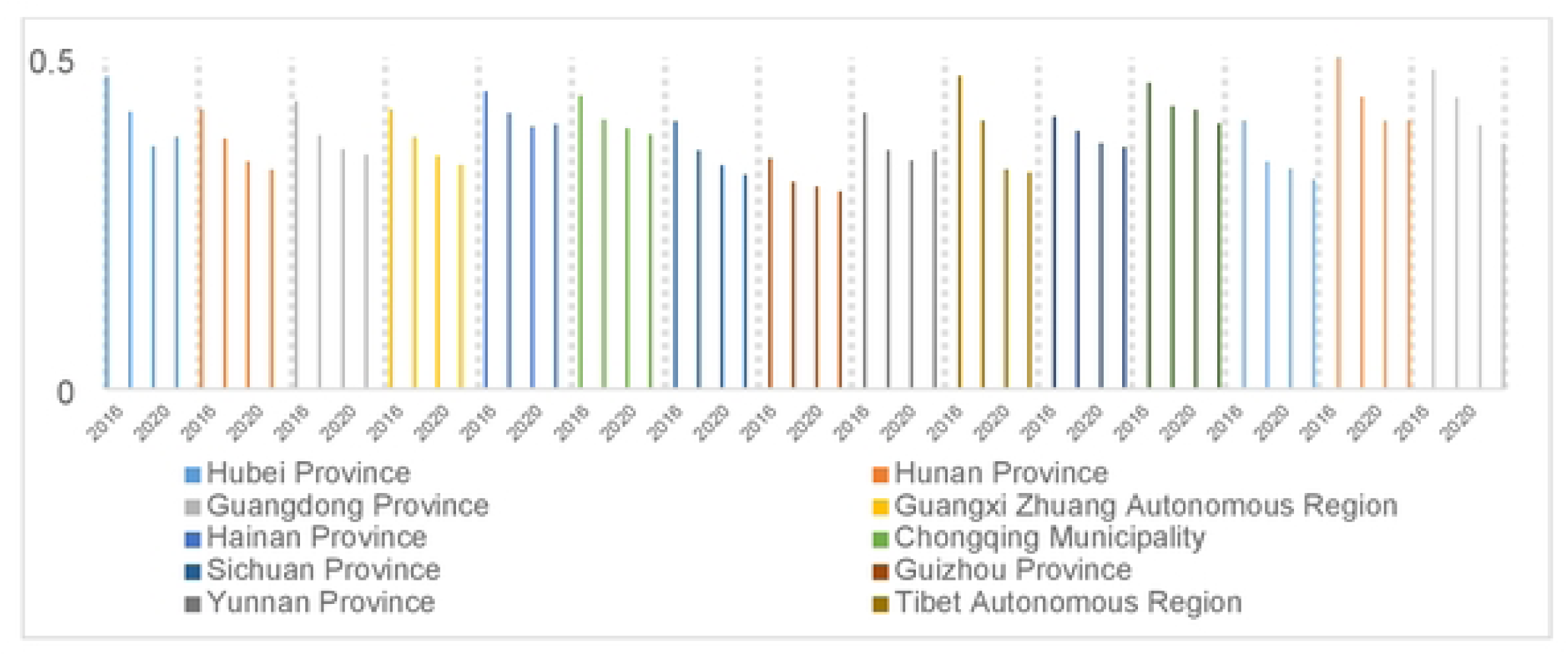
Average outpatient drug costs per visit as a percentage of total expenditures, 2016–2022.

**Figure 6.**
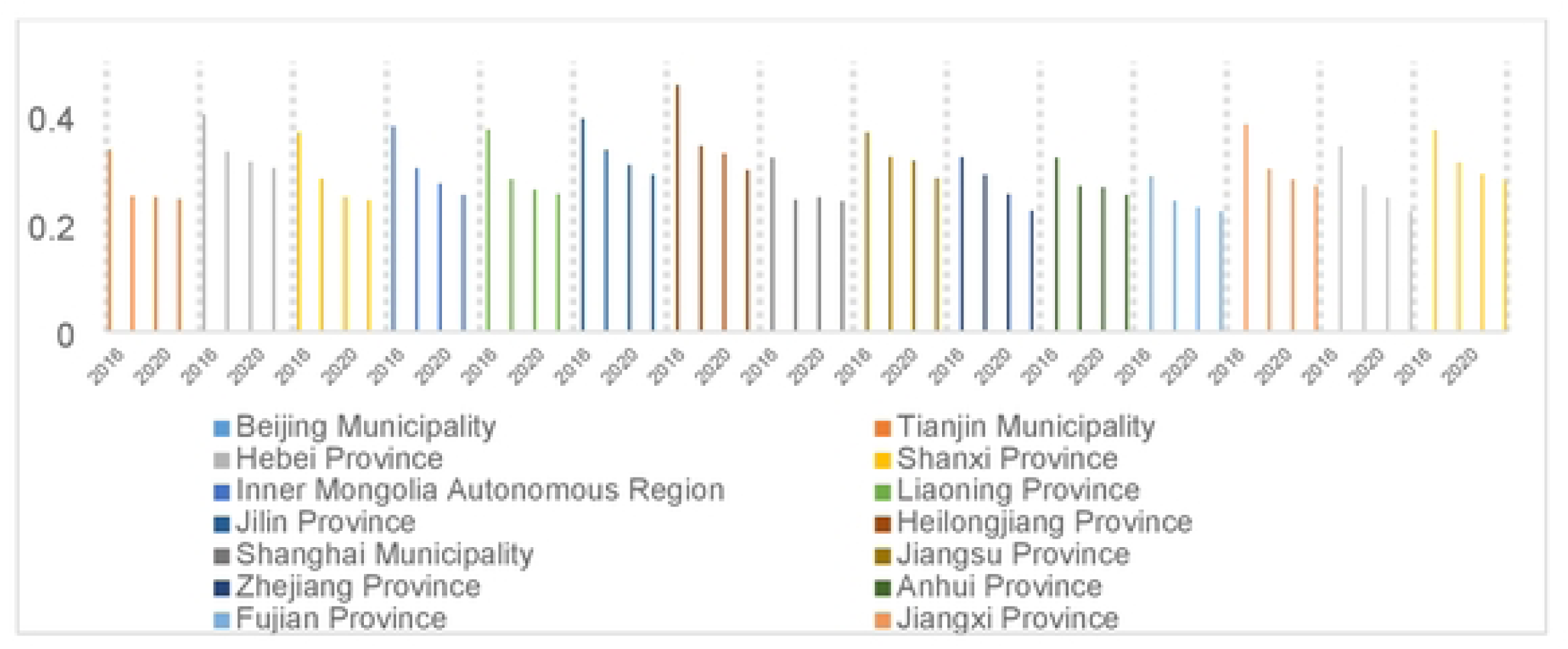
Average inpatient drug costs per hospitalization as a percentage of total expenditures, 2016–2022

**Figure 7.**
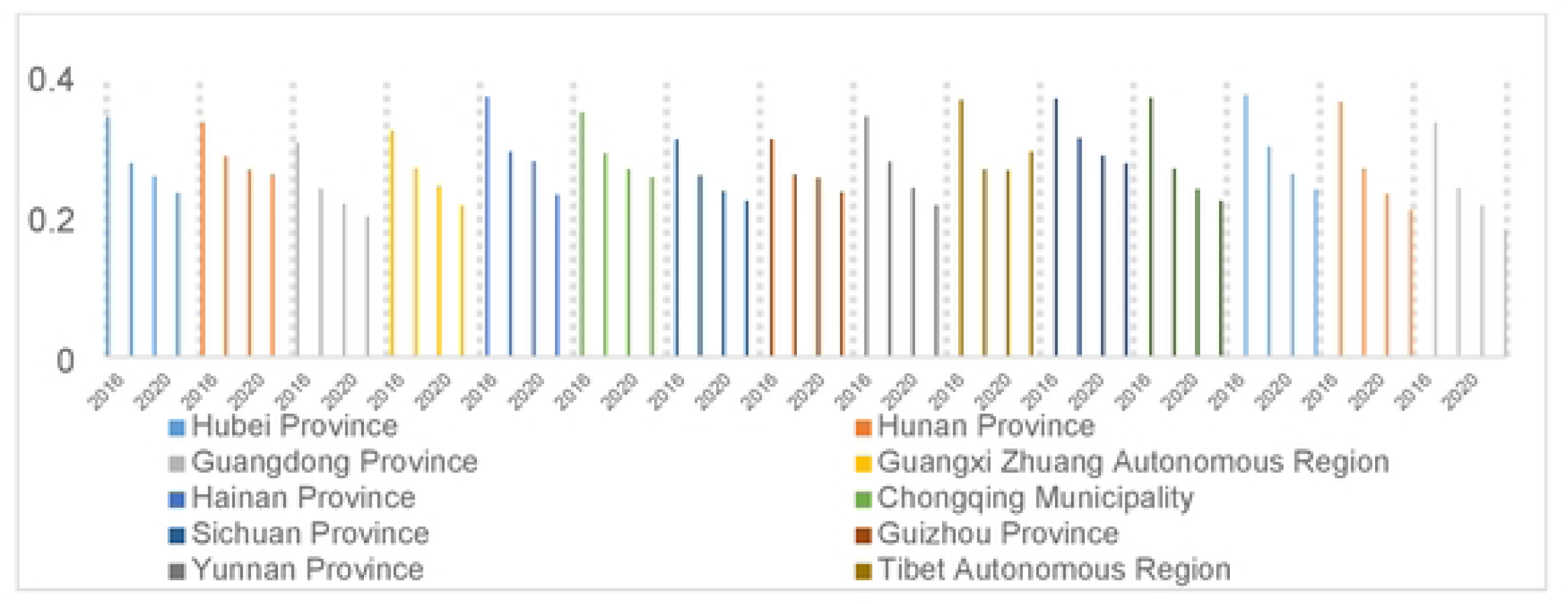
Average inpatient drug costs per hospitalization as a percentage of total expenditures, 2016–2022

As illustrated in Figures 4-7, the share of outpatient and inpatient drug expenditures in total medical costs exhibited a consistent year-on-year decline across provinces.

Table 12 confirms that the policy mix significantly reduced the per capita outpatient drug costs, demonstrating effective cost containment. However, the results of Model (4) indicated a positive association between inpatient drug expenditures and overall medical costs. This outcome was attributed to the expansion of insurance coverage for high-cost treatments, particularly for cancers and other severe conditions, under the national negotiation framework. This expanded inclusion has led to short-term surges in utilization and temporary increases in costs. Supporting evidence shows that the number of “nationally negotiated drugs” under the dual-channel mechanism grew from 221 in 2020 to 430 by 2024. For example, in Jiangsu Province, the total sales of such drugs rose from less than 4 billion yuan in 2020 to over 14.6 billion yuan by 2024, benefiting more than 3.6 million patients in 2020 and expanding to over 23 million patients by 2024.

**Table 12.** Pharmaceutical cost breakpoint regression results.

|  | (1)<br>Total average cost<br>per outpatient visit | (2)<br>Average cost of drugs<br>per outpatient visit | (3)<br>Total cost per<br>inpatient visit | (4)<br>Average cost of<br>drugs per inpatient<br>visit |
| --- | --- | --- | --- | --- |
| RD | -0.031***<br>(0.006) | -0.017**<br>(0.007) | -0.023**<br>(0.009) | 0.023**<br>(0.009) |
| Obs | 92097 | 92097 | 92097 | 92097 |

With respect to pharmaceutical cost reductions and their impact on health performance, the total medical expenditure captured under the effectiveness dimension can implicitly encompass drug costs. Therefore, this analysis isolated and examined pharmaceutical expenditures specifically in relation to performance outcomes. As shown in Table 13, outpatient and inpatient expenditures exhibited contrasting effects. The outpatient drug costs both per visit and in aggregate demonstrated significant negative correlations with health performance, confirming the effectiveness of outpatient cost control reforms. In contrast, inpatient care showed a different pattern. While per capita hospitalization costs were positively correlated with health outcomes, pharmaceutical expenditure within inpatient services showed statistically significant positive associations with health metrics. This apparent paradox suggested that while the systemic containment of hospitalization costs improved population-level health, targeted investment in pharmaceuticals remained essential for enhancing inpatient therapeutic outcomes. Accordingly, reforms to inpatient pharmaceutical management require a more refined approach. Rather than applying uniform cost-cutting measures, such reforms must aim to curb irrational drug use and inflated pricing while safeguarding access to clinically essential medications. Failure to account for the therapeutic value of specific drugs may compromise patient outcomes, especially among patients with serious or complex conditions that rely on critical drug therapies.

**Table 13.** Impact of outpatient pharmaceutical expenditures on population health performance.

|  | (1) | (2) | (3) | (4) |
| --- | --- | --- | --- | --- |
|  | Performance |  |  |  |
| Total average cost per outpatient visit | -0.047***<br>(0.009) |  |  |  |
| Average cost of drugs per outpatient visit |  | -0.024**<br>(0.012) |  |  |
| Total cost per inpatient visit |  |  | -0.049***<br>(0.011) |  |
| Average cost of drugs per inpatient visit |  |  |  | 0.034***<br>(0.007) |
| Obs | 92097 | 92097 | 92097 | 92097 |

### 6.2 Domestic pharmaceutical research and development

Using province-level pharmaceutical innovation data obtained from the Wind database and matched with CFPS data, this study incorporated province fixed effects to control for regional heterogeneity. R&D input and output indicators were measured using log-transformed R&D expenditures and the number of granted patents, respectively. Breakpoint regression was applied to evaluate the impact of policy implementation on pharmaceutical firms’ R&D activities, followed by fixed-effects models to examine how enterprise innovation could affect residents’ health performance.

#### 6.2.1 Firms’ R&D inputs

As shown in Table 14, Model (1) indicates that the implementation of medication payment policy increased provincial pharmaceutical R&D inputs by 5.7%, confirming that the policy has effectively strengthened innovation incentives. Models (2) through (4) revealed that each 1% increase in R&D investment led to a 0.027-point improvement in overall health performance, comprising a 0.008-point gain in performance and a 0.019-point gain in effectiveness, both statistically significant at the 1% level. These results suggest that pharmaceutical innovation indirectly enhances health outcomes through technological advancements. While medication cost reductions improved short-term efficiency, sustained improvements in population health required the long-term accumulation of innovation capacity.

#### 6.2.2 Firms’ R&D outputs

Model (5) in Table 14 indicated that the policy mix increased pharmaceutical R&D output by 7.9%, reflecting a substantial enhancement in innovation productivity. Models (6)–(8) further demonstrated that provincial R&D growth significantly improved population health performance. Specifically, each 1% increase in R&D output led to a 0.011-point rise in composite health scores, with effectiveness gains (0.008) nearly tripling performance improvements (0.003). This divergence is consistent with Models (2)–(4), suggesting that pharmaceutical innovation primarily enhances the efficiency of healthcare expenditure rather than directly boosting individual health outcomes.

**Table 14.** Mechanistic effects of firms’ R&D inputs and outputs.

| (1) | (2) | (3) | (4) | (5) | (6) | (7) | (8) |
| --- | --- | --- | --- | --- | --- | --- | --- |

|  | R&D<br>inputs | Health<br>perform<br>ance | perform<br>ance | effective<br>ness | R&D<br>outputs | Health<br>perform<br>ance | perform<br>ance | effective<br>ness |
| --- | --- | --- | --- | --- | --- | --- | --- | --- |
| RD | 0.057*<br>(0.033) |  |  |  | 0.079**<br>*<br>(0.026) |  |  |  |
| R&D<br>inputs |  | 0.027**<br>*<br>(0.002) | 0.008**<br>*<br>(0.001) | 0.019**<br>*<br>(0.001) |  |  |  |  |
| R&D<br>outputs |  |  |  |  |  | 0.016**<br>*<br>(0.002) | 0.004**<br>*<br>(0.001) | 0.012**<br>*<br>(0.001) |
| Obs | 92002 | 92002 | 92002 | 92002 | 92002 | 92002 | 92002 | 92002 |

### 6.3 Import and export of medicines

Using provincial pharmaceutical import-export data from China Customs (2016– 2022) that can be matched with household health survey indicators by province year, this study employed breakpoint regression to evaluate the impact of policy implementation on the medicine trade. Fixed-effects models were subsequently applied to assess trade’s influence on health performance outcomes.

#### 6.3.1 Drug import amount

Model (1) in Table 15 confirmed that pharmaceutical imports declined after 2019, largely due to procurement and negotiation policies. Reduced import prices discouraged foreign suppliers, whereas the dominance of domestic bids further lowered reliance on imported medicines. Models (2)–(4) showed that each 1% increase in import volume was associated with a marginal improvement of 0.003 units in health performance, driven primarily by gains in effectiveness. This reflects the concentration of imports in high-efficacy therapies, which can enhance resource allocation. However, import dependence remains limited to specific conditions and population groups. Barriers such as reimbursement constraints and affordability issues limited broader health gains, suggesting that while import suppression could promote domestic substitution, it may also moderately restrict the scope of population-level health improvements.

#### 6.3.2 Export value of pharmaceuticals

Table 15 results reveal a significant increase in pharmaceutical exports following policy intervention. The centralized procurement incentivized domestic firms to improve cost control and enhance international price competitiveness. Concurrently, national negotiations facilitated the inclusion of innovative domestic drugs in health insurance coverage, thereby releasing cash flows for overseas market expansion. Models (6)–(8) indicated that each 1% increase in export value contributed to a 0.022-unit increase in overall health performance, with 0.007 attributed to improved health outcomes and 0.015 attributed to enhanced expenditure efficiency. These findings demonstrated that medication payment policies promoted export-driven pharmaceutical innovation, improved drug quality for domestic use, and generated economies of scale that reduced production costs. This exerted an indirect downward pressure on domestic healthcare expenditure. In summary, the policy combination supported population health through two channels: improving drug quality and optimizing cost efficiency.

**Table 15.**
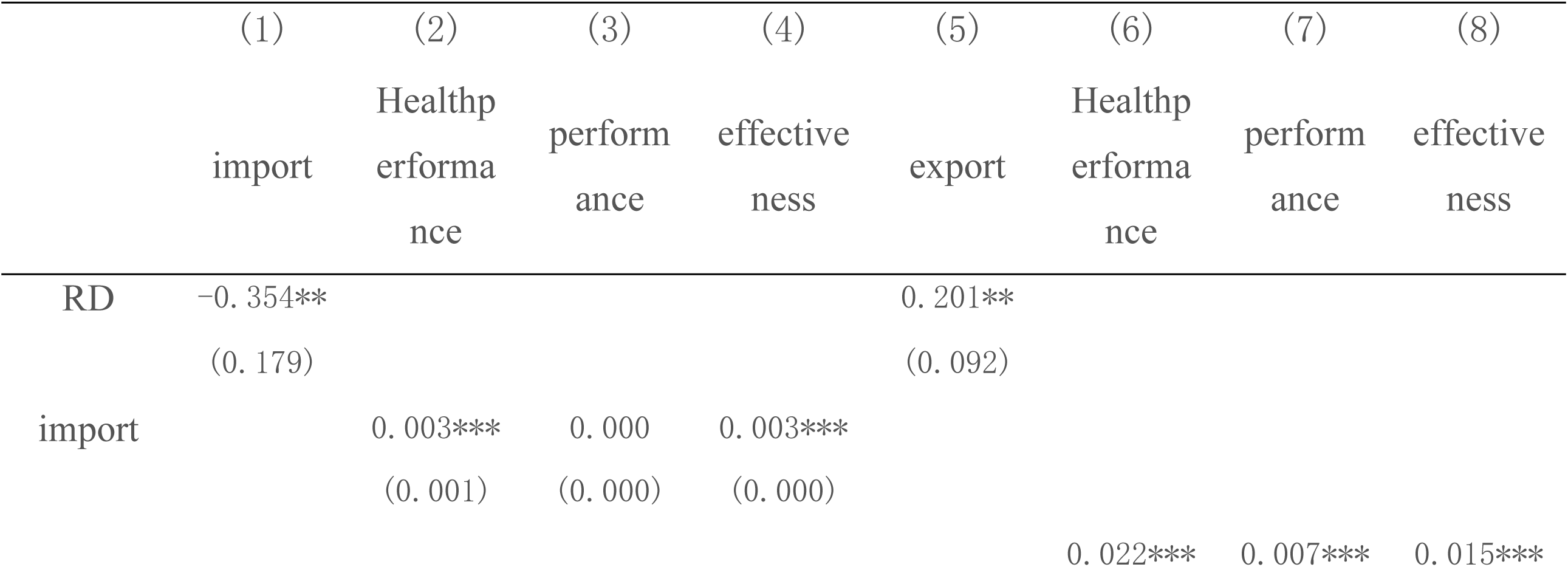
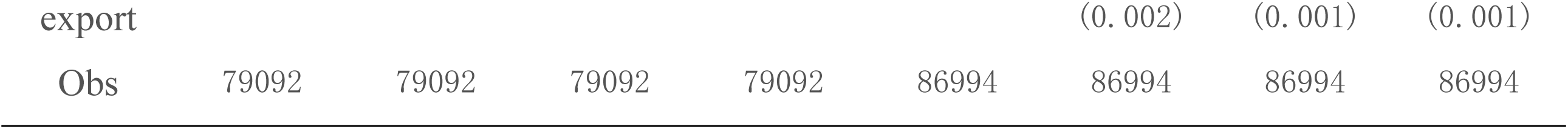
Effects of mechanisms for import and export of pharmaceuticals.

In summary, Hypothesis H2 was verified.

## 7. Research Conclusion and Policy Recommendations

### 7.1 Research Conclusions

This study constructed a two-dimensional performance evaluation framework based on “health outcomes” and “medical expenditures” and systematically assessed the impact of medication payment and management policies on population health performance. The analysis was performed based on breakpoint regression models and leveraged CFPS microdata along with pharmaceutical import-export statistics. The key research findings are summarized as follows:

7.1.1 The Medicare payment management policy mix significantly improves population health performance across both performance and efficiency dimensions. After controlling for provincial fixed effects, time trends, and relevant covariates, the policy was found to increase the population health outcome index by 1.1% and enhance healthcare spending efficiency by 2.4%, with the results validated through robustness tests.

7.1.2 Notable regional heterogeneity was observed. In Eastern China, the policy effects were statistically insignificant, likely due to saturation in mature healthcare markets. In Central China, the policy primarily improved healthcare expenditure efficiency (i.e., enhancing effectiveness but not performance). Western China achieved significant gains in both health outcomes and expenditure efficiency, underscoring greater policy responsiveness in underdeveloped regions.

7.1.3 Mechanism analysis revealed that the policy enhanced residents’ net health gains through three main pathways: reducing out-of-pocket medication costs, incentivizing pharmaceutical R&D investment and innovation, and optimizing medicine trade structures. Improvements were realized by curbing imports and promoting exports, with effectiveness-related gains substantially exceeding performance-related improvements.

### 7.2 Policy Recommendations

#### 7.2.1 Dynamic adjustment of the division between volume procurement and national negotiations to enhance drug classification management

The classification of volume-based procurement and national drug negotiation should be refined dynamically. Volume procurement is advised for mature pharmaceuticals (consistency-evaluated generics and off-patent originators), where competitive bidding mechanisms can compress costs and secure basic drug access for vulnerable populations. National negotiations should prioritize clinically urgent, high-value, and innovative therapies to expand coverage and improve accessibility and health outcomes.

Different payment mechanisms are also recommended. For essential therapeutics, “supply-guaranteed negotiation plus installment” strategies serve to prevent price suppression-induced shortages. For auxiliary drugs, the use of negative lists with stringent prescription and dosage monitoring supports more efficient resource allocation. For exploratory drugs, clinical data platforms can be created with efficacy benchmarks, enabling automatic delisting of underperforming drugs to ensure catalog quality and value.

#### 7.2.2 Establishing a quality-benefit two-dimensional evaluation system and enhancing full-cycle medicine regulation

For collectively procured drugs, a quality tracing mechanism can be implemented and the frequency and scope of pharmacological inspections can be enhanced through joint regulatory supervision. Enterprises with repeated inspection failures should be blacklisted. For innovative drugs under insurance coverage, efficacy monitoring should be conducted every 3–5 years using hospital EMRs and insurance settlement data to assess health outcomes. Based on these evaluations, renegotiation or withdrawal of underperforming products should be initiated.

#### 7.2.3 Regionally differentiated supply strategies to address structural imbalances in medical resources

Pilot intra-/extra-directory coordinated payment mechanisms in eastern China to help high-income populations access non-listed innovative drugs through commercial insurance tailored to differentiated needs. In the central and western regions, fiscal transfers should be increased to expand the scope of collective procurement, ensure the supply of basic medicines, and meet rising demand for rational medication.

#### 7.2.4 Establishing a full-chain policy system to accelerate industrial transformation

A full-chain policy system should be established to drive pharmaceutical industrial transformation. First, R&D risk-sharing can be promoted through national funds targeting major disease drugs, while expanding R&D tax deductions to reduce innovation costs. Second, market access can be accelerated by creating fast-track approval pathways for breakthrough therapies, exempting routine pharmacoeconomic assessments, and completing insurance negotiations within six months to accelerate patient benefits. Third, tertiary hospitals must allocate dedicated budgets for clinically validated innovative drugs, thereby removing barriers to access within the hospital systems. This integrated policy design is expected to reduce R&D and market-entry uncertainty, redirect corporate investment toward high-value clinical areas, improve health efficiency, and ultimately transition China’s pharmaceutical sector from generic production to innovation-driven development.

## Data Availability

The data that support the findings of this study are derived from the China Family Panel Studies (CFPS). The CFPS data are collected and distributed by the Institute of Social Science Survey (ISSS) at Peking University. The original data used in this study can be accessed upon application through the official CFPS website (https://www.isss.pku.edu.cn/cfps/).

